# Social inequalities in child development: Analysis of Low-Birth-Weight trends in Brazil, 2010-2020

**DOI:** 10.1101/2022.11.29.22282908

**Authors:** Audêncio Victor, Italo Wesley Oliveira Aguiar, Renzo Flores-Ortiz, Manuel Mahoche, Ana Raquel Manuel Gotine, Ila Falcão, Melsequisete Daniel Vasco, Andrêa Ferreira, Mark Omenka, José Leopoldo Ferreira Antunes, Patrícia H. Rondo

## Abstract

**Introduction:** Globally, low birth weight (LBW) is prevalent in low-income countries. Although the economic assessment of interventions to reduce this burden is essential to guide health policies, research that illustrates the magnitude of LBW by country and region as a way to support the design of public policies is still relatively scarce.

**Objective:** To analyze the temporal trend of fetal growth of newborns in Brazil, in the period from 2010 to 2020.

**Methods:** A time series study, whose data source was the Live Births Information System (SINASC), of the Department of Information and Informatics of the Unified Health System (DATASUS), linked to the Ministry of Health, Brazil. The Prais-Winsten linear model was applied to analyse the annual proportions of LBW. The annual percentage changes (APC) and its respective 95% confidence intervals (95%CI) were calculated. To visualize the dynamics of evolution in each Federation Unit (FU), prevalence rate averages of LBW were calculated and displayed on thematic maps.

**Results:** Between 2010 and 2020, there was a trend toward stabilization of the increasing proportions of low birth weight in the North, Northeast and Centre-West regions. In Brazil and the other regions these tendencies remained stable.

**Conclusion:** To improve the living conditions of the population, public policies and actions aimed at reducing social inequalities and inequity is health are necessary, such as strengthening the Unified Health System (SUS), income transfer programs, quota policies for vulnerable groups, and gender to improve access to education for women and the labour sector.

**WHAT IS ALREADY KNOWN ON THIS TOPIC:**

- In Brazil, health social inequalities have a historical explanation due to the socio-economic and political system that encourages a competitive economic development model.
- Improved access to education and health care has reduced the risk of low birth weight (LBW) in all Brazilian regions in recent years.
- Differences in trends in LBW rates and associated factors within and between regions of Brazil.

**WHAT THIS STUDY ADDS:**

- The study provides relevant information on the trend of LBW rates in recent years and makes a comparison between regions and the country as a whole.
- There were differences in the trend of LBW between Brazilian regions, with an increasing trend in the North, Northeast and Central-West regions, but on the other hand, the South and Southeast regions remain stationary.
- Stationary trend in the prevalence of BPN in Brazil in recent years (2010-2020), findings that should be associated with the improvement in the living conditions of the population.

**HOW THIS STUDY MIGHT AFFECT RESEARCH, PRACTICE OR POLICY:**

- It can be taken into account when building public policies aimed at improving the living conditions of the population public policies and actions aimed at reducing health inequalities and inequities are needed.

## Introduction

Birth weight is the first weight recorded after birth and an important indicator for health outcomes in later periods of the child’s favorable life (1–7), LBW is defined by the World Health Organization (WHO) as birth weight below 2,500g, regardless of gestational age (8). LBW is an important public health indicator reflecting maternal and neonatal health both from a nutritional, economic and health care nutrition perspective, as newborns (NBs) with LBW are at higher risk of morbidity and mortality from death and illness soon after birth and noncommunicable diseases throughout life (3,4,7,9).

It has been proven that low birth weight NBs are 20 times more predisposed to develop complications and die compared to normal weight NBs (Wardlaw, 2004). LBW predisposes NBs to a potential risk of cognitive deficits, motor delays, cerebral palsy and other behavioural and psychological problems (6,10,11). Also, household costs as well as health system costs could be saved by reducing the burden of LBW (12). Despite the great concern of governments and states about reducing LBW, there are still scarce research that assesses the temporal trends of this problem. A study evaluating trends in prematurity, LBW, and intrauterine growth restriction in three birth cohorts from 1982, 1993, and 2004 in Pelotas, Brazil, observed a slight increase in the prevalence of LBW from 9% to 10%. Intrauterine growth restriction decreased from 14.8% in 1982 to 9.4% in 1993, and subsequently increased to 12% in 2004, while preterm births increased sharply, from 6.3% in 1982 to 14.7% in 2004 (13).

Improved access to education and health care reduced the risk of LBW in all Brazilian regions between 1996 and 2011, in a study that analysed the trend of LBW and its determinants in Brazilian capitals (14). The same study, highlighted differences in trends in LBW rates and associated factors within and between regions looking at the stages of demographic, epidemiological, and developmental transition in Brazil (14). To understand the variation of LBW over time, as well as compare the temporality of LBW (15,16), looking at the different factors that may be potential determinants of this phenomenon, we sought to analyse the temporal trends for LBW from 2000 to 2020.

### Methods

Ecological study with time series analysis on fetal growth involving proportion of live births with LBW in Brazil, referring to the period from 2010 to 2020. The study data were obtained from the Live Births Information System (SINASC) of the Department of Information and Informatics of the Unified Health System (DATASUS) of the Ministry of Health, Brazil through the Platform for Data Science Applied to Health (PCDaS/Icict/Fiocruz) (17).

### Study variables

The outcome LBW was defined by the dichotomous classification of the variable birth weight: LBW (< 2500g) and normal weight (> 2500g). The LBW prevalence rate was calculated using the number of NBs with LBW divided by the total population of NBs in the place and year, multiplied by 100. The rates were calculated according to the Brazilian macro-regions.

### Statistical Analysis

The Prais-Winsten linear regression model, which considers serial autocorrelation, i.e. the dependence of a serial measure on its own values in previous periods, was used to analyse the temporal trend of annual fetal growth rates. The annual percentage changes (APC) and its respective 95% confidence intervals (95%CI) were calculated. To visualise the dynamics of evolution in each region, LBW prevalence rate averages were calculated and displayed on thematic maps by QGIS 2.18 software.

The trend of LBW prevalence was interpreted as increasing (p<0.05 and positive beta), decreasing (p<0.05 and negative beta) and stable (p≥0.05), as designed by Antunes & Cardoso (2015). The exploration of the explanatory variables and the analysis of time series were performed by the program Stata version 14.0 (Stata Corp., College Station, United States).

## Results

A total number of 31,887,329 women from all Federative Units of Brazil were analysed over the years of the study. Among the participants in the study, the largest proportion is from the Southeast region. And 9.5% of the records were for the year 2015. Among them, 49.6% of women were between 20 and 29 years old, with more than half (75.5%) having between 8 and 12 years of schooling. Their newborns were (58.8%) male and non-white (Table 1).

**Table 1.** Maternal and newborn sociodemographic characteristics in Brazil, 2010-2020

| Variables | n | % |
| --- | --- | --- |
| **Regions** |  |  |
| North | 3.439.187 | 10.8 |
| Northeast | 9.051.960 | 28.4 |
| Southeast | 12.528.721 | 39.3 |
| South | 4.265.927 | 13.4 |
| Central West | 2.601.534 | 8.2 |
| Brazil | 31.887.329 | 100 |
| **Year of birth of the baby** |  |  |
| 2010 | 2.861.868 | 8.97 |
| 2011 | 2.913.160 | 9.14 |
| 2012 | 2.905.789 | 9.11 |
| 2013 | 2.904.027 | 9.11 |
| 2014 | 2.979.259 | 9.34 |
| 2015 | 3.017.668 | 9.46 |
| 2016 | 2.857.800 | 8.96 |
| 2017 | 2.923.535 | 9.17 |
| 2018 | 2.944.932 | 9.24 |
| 2019 | 2.849.146 | 8.94 |
| 2020 | 2.730.145 | 8.56 |
| **Age category (Year)** |  |  |
| < 20 | 5.583.175 | 17.51 |
| 20-29 | 15.825.989 | 49.63 |
| >= 30 | 10.477.889 | 32.86 |
| **Schooling (years)** |  |  |
| None | 205.041 | 0.64 |
| 1 a 3 | 1.020.054 | 3.20 |
| 4 a 7 | 6.095.600 | 19.12 |
| 8 a 11 | 18.184.596 | 57.03 |
| 12 | 5.883.787 | 18.45 |
| Ignored | 498.251 | 1.56 |
| **Sex of newborn** |  |  |
| Male | 16.329.184 | 51.21 |
| Female | 15.552.623 | 48.77 |
| Ignored | 5.522 | 0.02 |
| **Race/ethnicity of newborn** |  |  |
| White | 11.700.134 | 36.69 |
| Black | 1.603.373 | 5.03 |
| Yellow | 119.209 | 0.37 |
| Brown | 17.014.744 | 53.36 |
| Indigenous | 251.422 | 0.79 |
| Ignored | 1.198.447 | 3.76 |

From 2010 to 2020, the yearly percentage variation of the proportion of LBW in all Brazilian Federal Units (UF) was equivalent to 0.18%. The lowest proportion of LBW was -0.05% in the Southeast, while the North, Northeast, and Center-West had an increasing trend in the proportion of LBW while in the South and Southeast a stationary trend was observed (Table 2).

**Table 2.**
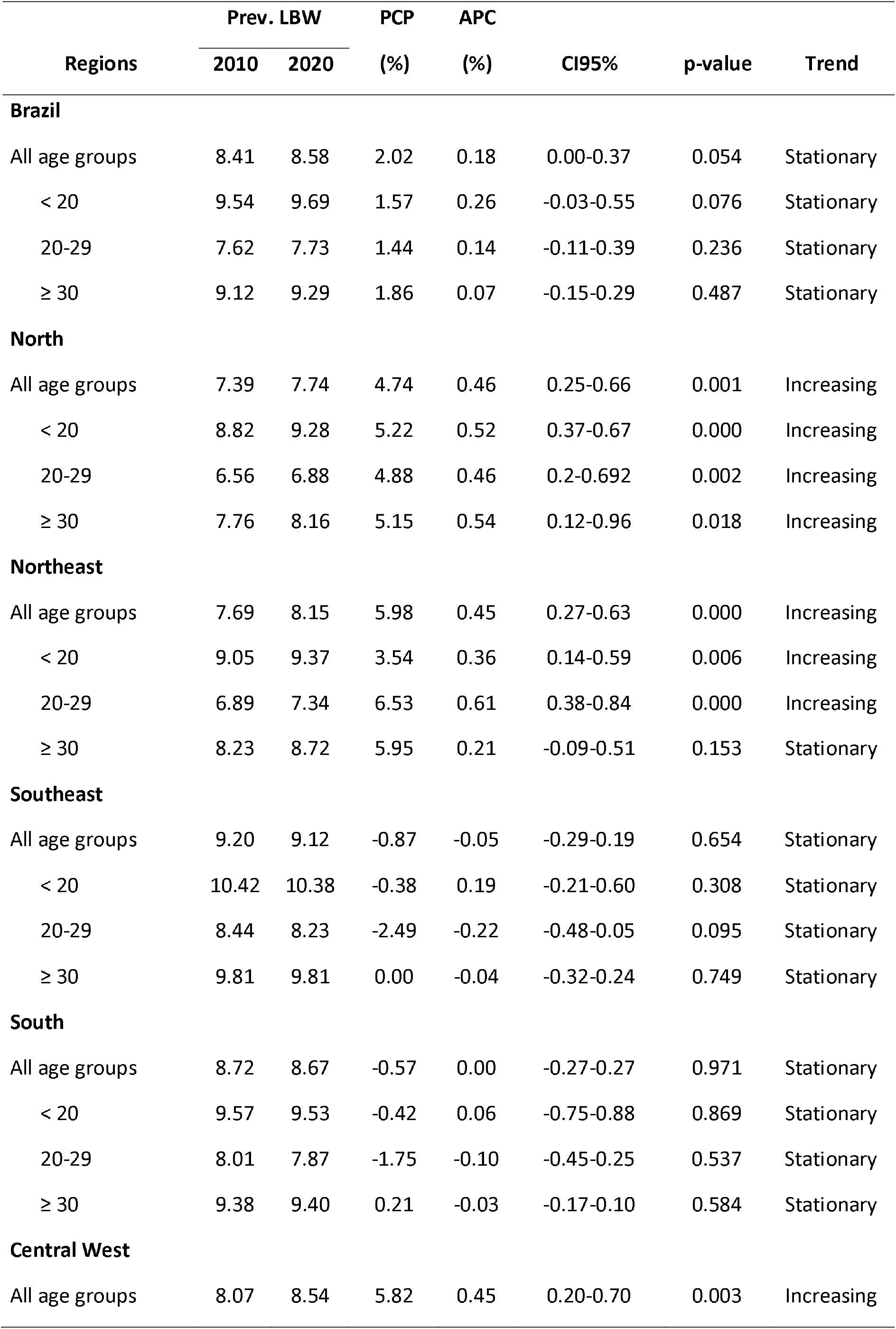
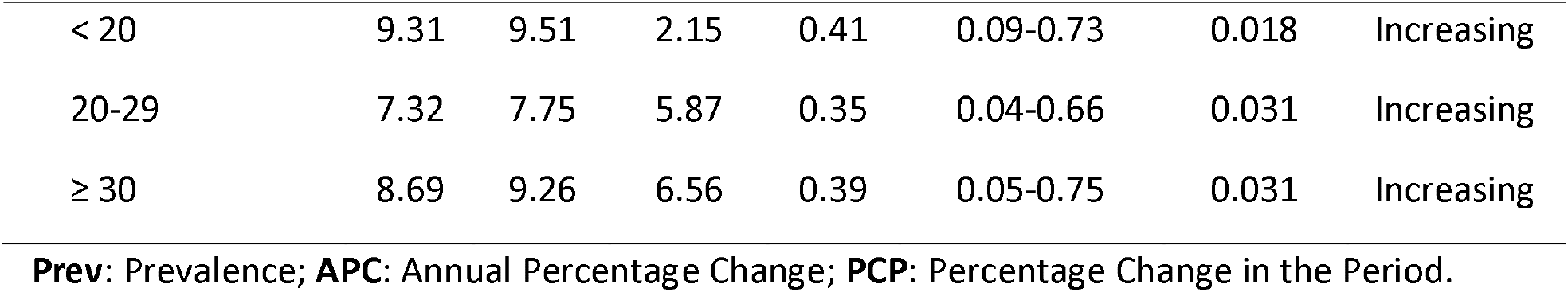
LBW Prevalence rate and yearly variation according to maternal age, Brazil, 2010-2020

| Regions | Prev. LBW |  | PCP | APC | CI95% | p-value | Trend |
| --- | --- | --- | --- | --- | --- | --- | --- |
|  | 2010 | 2020 | (%) | (%) |  |  |  |
| Brazil |  |  |  |  |  |  |  |
| All age groups | 8.41 | 8.58 | 2.02 | 0.18 | 0.00-0.37 | 0.054 | Stationary |
| < 20 | 9.54 | 9.69 | 1.57 | 0.26 | -0.03-0.55 | 0.076 | Stationary |
| 20-29 | 7.62 | 7.73 | 1.44 | 0.14 | -0.11-0.39 | 0.236 | Stationary |
| ≥ 30 | 9.12 | 9.29 | 1.86 | 0.07 | -0.15-0.29 | 0.487 | Stationary |
| North |  |  |  |  |  |  |  |
| All age groups | 7.39 | 7.74 | 4.74 | 0.46 | 0.25-0.66 | 0.001 | Increasing |
| < 20 | 8.82 | 9.28 | 5.22 | 0.52 | 0.37-0.67 | 0.000 | Increasing |
| 20-29 | 6.56 | 6.88 | 4.88 | 0.46 | 0.2-0.692 | 0.002 | Increasing |
| ≥ 30 | 7.76 | 8.16 | 5.15 | 0.54 | 0.12-0.96 | 0.018 | Increasing |
| Northeast |  |  |  |  |  |  |  |
| All age groups | 7.69 | 8.15 | 5.98 | 0.45 | 0.27-0.63 | 0.000 | Increasing |
| < 20 | 9.05 | 9.37 | 3.54 | 0.36 | 0.14-0.59 | 0.006 | Increasing |
| 20-29 | 6.89 | 7.34 | 6.53 | 0.61 | 0.38-0.84 | 0.000 | Increasing |
| ≥ 30 | 8.23 | 8.72 | 5.95 | 0.21 | -0.09-0.51 | 0.153 | Stationary |
| Southeast |  |  |  |  |  |  |  |
| All age groups | 9.20 | 9.12 | -0.87 | -0.05 | -0.29-0.19 | 0.654 | Stationary |
| < 20 | 10.42 | 10.38 | -0.38 | 0.19 | -0.21-0.60 | 0.308 | Stationary |
| 20-29 | 8.44 | 8.23 | -2.49 | -0.22 | -0.48-0.05 | 0.095 | Stationary |
| ≥ 30 | 9.81 | 9.81 | 0.00 | -0.04 | -0.32-0.24 | 0.749 | Stationary |
| South |  |  |  |  |  |  |  |
| All age groups | 8.72 | 8.67 | -0.57 | 0.00 | -0.27-0.27 | 0.971 | Stationary |
| < 20 | 9.57 | 9.53 | -0.42 | 0.06 | -0.75-0.88 | 0.869 | Stationary |
| 20-29 | 8.01 | 7.87 | -1.75 | -0.10 | -0.45-0.25 | 0.537 | Stationary |
| ≥ 30 | 9.38 | 9.40 | 0.21 | -0.03 | -0.17-0.10 | 0.584 | Stationary |
| Central West |  |  |  |  |  |  |  |
| All age groups | 8.07 | 8.54 | 5.82 | 0.45 | 0.20-0.70 | 0.003 | Increasing |
| < 20 | 9.31 | 9.51 | 2.15 | 0.41 | 0.09-0.73 | 0.018 | Increasing |
| 20-29 | 7.32 | 7.75 | 5.87 | 0.35 | 0.04-0.66 | 0.031 | Increasing |
| ≥ 30 | 8.69 | 9.26 | 6.56 | 0.39 | 0.05-0.75 | 0.031 | Increasing |

**Prev:** Prevalence; **APC:** Annual Percentage Change; **PCP:** Percentage Change in the Period.

Analysis of the results presented in Graph 1. shows that there is a low APC in the proportions of LBW in Brazil between 2010 and 2020 (0.18%). But the disaggregated results show that the Northeast, Midwest, and North regions recorded a slight increase in the prevalence of LBW.

Analysing how the distribution of the proportions of LBW has evolved in the 10 years for all the Federal States, we can observe a stabilisation of the trends in general, although there is variability of behaviour within and between the regions (Figure 2), as well as the APV, of 0.46% in the Southern region and 0.45% in the North-East and Central-Western regions, respectively.

**Figure 1.**
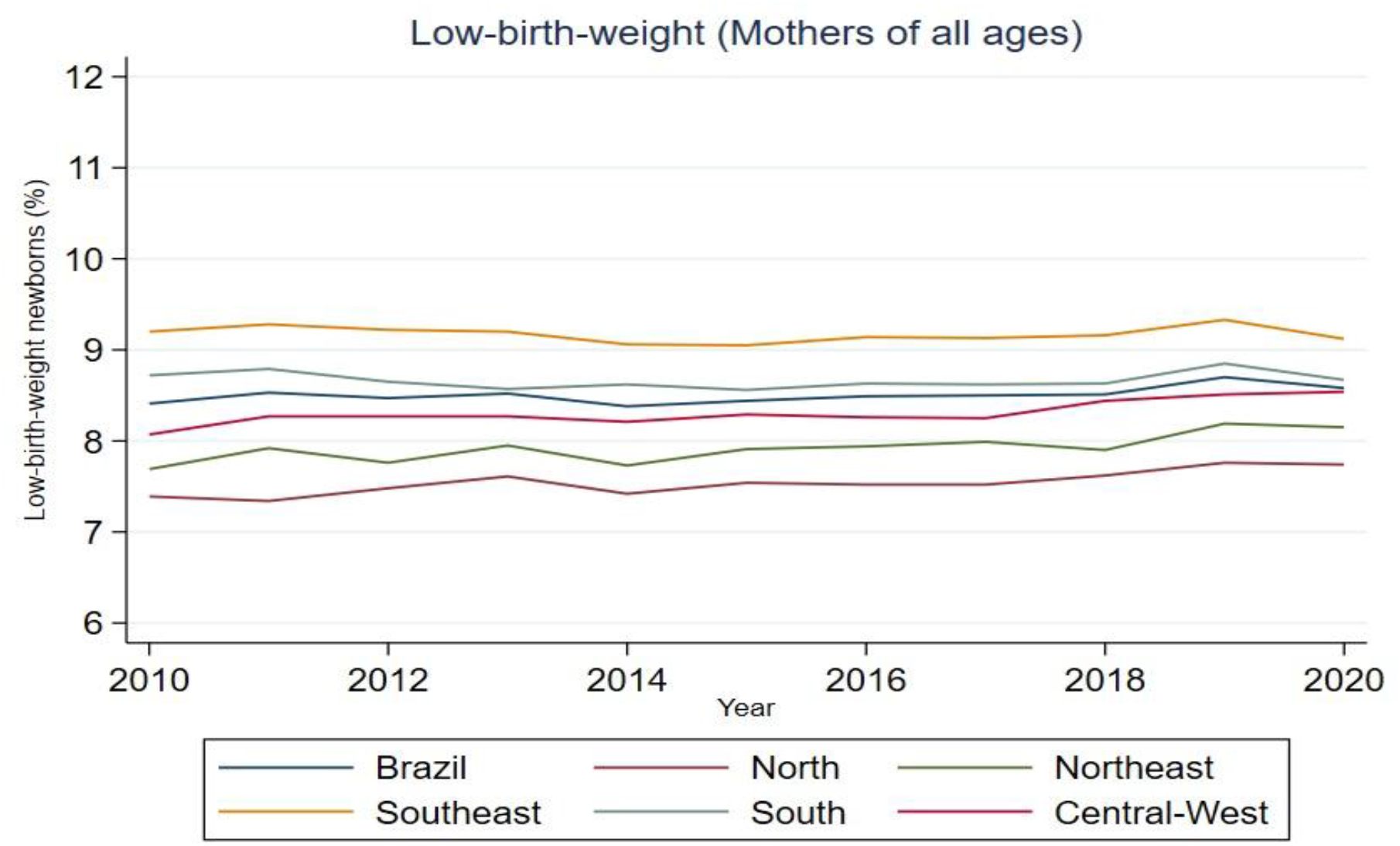
The proportion of LBW (all Brazilian mothers) according to year of notification.

**Figure 2.**
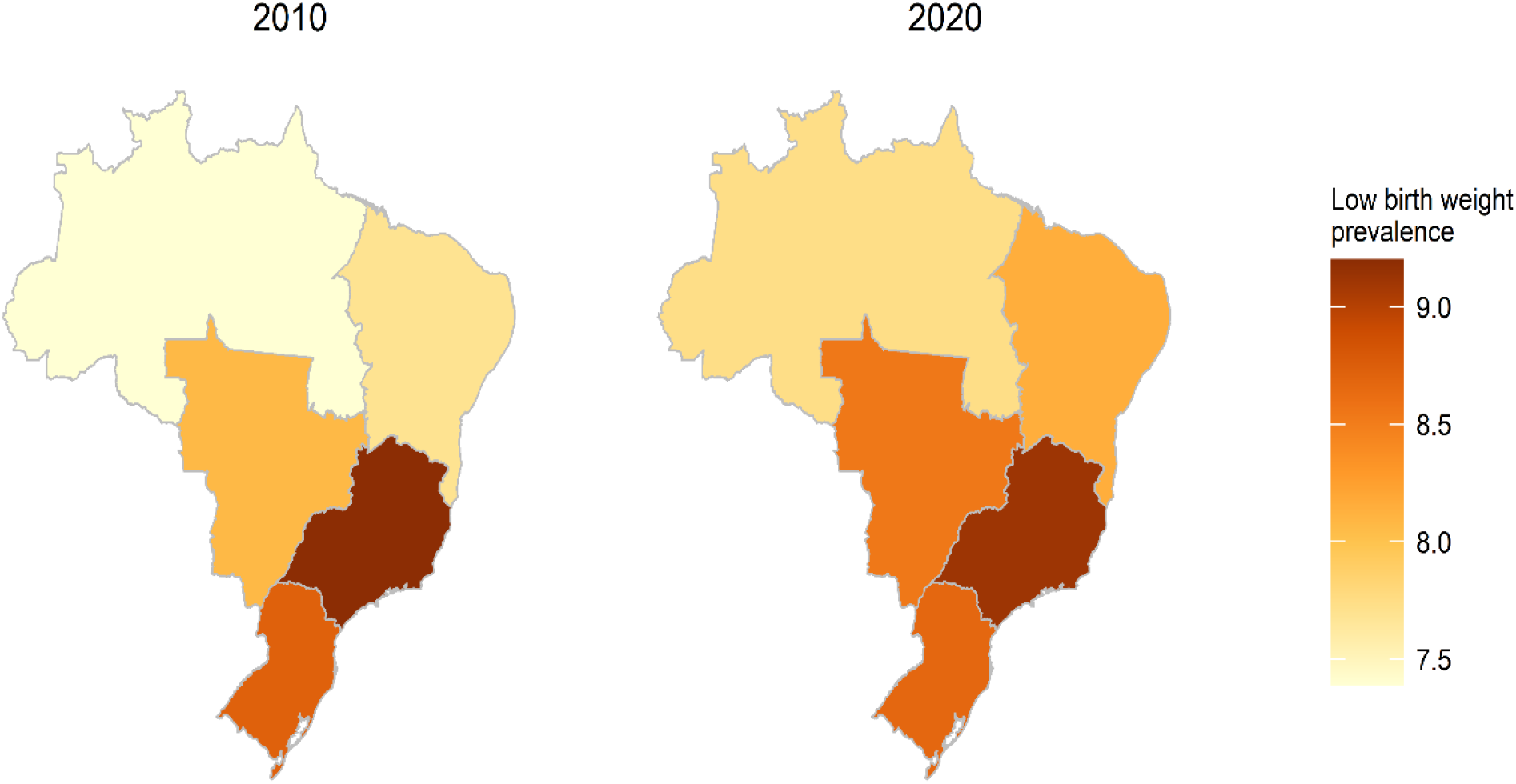
Prevalence of LBW in the Brazilian macro-regions in 2010 and 2020

## Discussion

Our findings show a stationary trend in the prevalence of LBW in Brazil in recent years. These findings may be associated with the improvement in access to health services, in the quality and quantity of prenatal consultations, in the socioeconomic conditions concretely in women’s education and massive entry of women in the labour market in general (14).

Despite these advances, point prevalence rates among the South, Southeast and Centre-West regions showed high prevalence rates of LBW. Therefore, we highlight that these results should be interpreted carefully, considering that these regions are part of the most developed ones in the country and with better health levels. Similar findings were observed in the study by Lima et al (18), which calls this phenomenon the *low birth weight paradox*, because the highest percentages of LBW are observed in the regions with better socioeconomic status (18). Another study that aimed to evaluate the temporal trend of LBW in Brazil from 1996 to 1999 showed that the states of Minas Gerais, Rio de Janeiro, São Paulo and Rio Grande do Sul had the highest rates of LBW among all the states (19).

Experts in the area attribute this phenomenon to the fragility of SINASC coverage in the regions with the worst levels of health. However, the same cannot be said today, since the coverage of this system is adequate and the quality of the information has improved, being considered a relevant source of data for health research and evaluation. The use of the data available in this system has been essential for the surveillance and monitoring of health inequalities and the quality of care (20– 22).

On the other hand, these results may be associated with the improvement in the quality of diagnosis and care to pregnant women that has been enabling the induction of deliveries including caesarean sections, which from the public health point of view, is a serious problem. Thus, there is an increase in preterm births and in the proportions of LBW, despite the reduction in infant mortality, especially in regions that offer better perinatal health services (23). However, inadequate care and precarious access to perinatal technology in disadvantaged areas result in deaths of live births soon after birth, with consequent underreporting or registration with misclassification, thus reducing the proportions of prematurity and low birth weight births (23,24).

But the findings related to the temporal trend of LBW show variations between Brazilian regions, with an increasing trend in the proportions of LBW in the North, Northeast and Center-West regions, but on the other hand in the South and Southeast regions the trends remain stationary regardless of the maternal age group. Therefore, it is convenient to remember that this spatial distribution of the rates of LBW in the states is related to space and shows the social and health inequality experienced in the Brazilian territory. Historically, in the North and Northeast regions, the offer and access to health services are restricted (access to assistance and adequate prenatal ICU beds). On the other hand, these are the regions, where pregnant women mostly experience socio-economically disadvantaged situations, this clearly exposes Brazil’s social problem situation as a precursor of health inequities, which culminates in possible problems during pregnancy, during delivery, in the first days of life, including the development of cognitive skills (25). In Brazil, research conducted in different regions indicates that mothers belonging to more socially vulnerable groups receive lower quality prenatal care (26,27).

Brazil is an extremely unequal country at all three levels (Federal, State and Municipal). Possible explanations include the historical legacy of a socio-economic and political system that encourages an economic development model of competitiveness to the detriment of integrality. Health is no different, as our results point out, where regionalization deeply affects the unequal distribution of resources, exposed through the setback in the project of universalization of access to the SUS (28).

In this context, it is worth noting the political phenomenon of dismantling the project of universal free SUS as well as limitations of other social policies imposed by EC 95/2016 (29), and the financialization of social and health policies (30). This culminates on the one hand with the extinction and replacement of policies with quite beneficial effects known as the Stork Network for the Maternal and Child Care Network (RAMI), in the policy of care for pregnant women, and on the other hand with the discontinuity of universal policies, and the commodification of health services (30), including those at the primary care level, thus widening the gap for access and coverage in the supply of services and limitation of health rights. According to these results, Brazil remains challenged to achieve the targets in the second sustainable development goal (SDG2), which consists of reducing the incidence of LBW and contributing to the reduction of infant mortality (31).

### Strengths and limitations

The quality and reliability of secondary data. In this regard, data were obtained from government sources, such as health information systems, which are known to have high quality standards.

## Conclusion

The findings show a spatial and temporal distribution of the rates of LBW, which are unevenly distributed in the Brazilian territory. The inequality of LBW reflects the health and socioeconomic conditions of the Brazilian states, which were historically marked by public policies that disrespect the social and spatial dynamics, in addition to the capitalist dynamics that leaves to the margin of economic flows the areas that do not offer infrastructure and qualified labour, reflecting on the health and life of people. After verifying that the more developed regions of the country have higher rates of LBW, we then conclude that the presence of health services and their use by the population decrease the numbers of infant mortality and increase the rates of LBW. Therefore, in order to improve the living conditions of the population, public policies and actions aimed at reducing inequalities and inequities in health are necessary, such as strengthening the Unified Health System (SUS), income transfer programs, quota policies for vulnerable groups, gender for access to education and the labour market to improve living conditions.

## Data Availability

The study data were obtained from the Live Births Information System (SINASC) of the Department of Information and Informatics of the Unified Health System (DATASUS) of the Ministry of Health, Brazil through the Platform for Data Science Applied to Health (PCDaS/Icict/Fiocruz)

https://pcdas.icict.fiocruz.br/criar-conta/?voltar=https://pcdas.icict.fiocruz.br/conjunto-de-dados/sistema-de-informacao-sobre-nascidos-vivos/documentacao/

## Acknowledgments

**Acknowledgments** not applicable.

## Financial support

Not applicable.

## Conflict of Interest

The authors declare no conflicts of interest

## Authorship

The authors’ contributions were as follows: A.V., A.R.M.G. and M.M. designed the study: A.V., I.W.O.A. and R.F.O. collected data and constructed the database: A.V, I.W.O.A, and L.A. outlined the analytical strategy: A.V., L.A, R.F.O and I.OW.A. performed the statistical analyses, interpreted the results and drafted the manuscript: M.M, P.R, A.R.M.G, A.F, L.A and M.D.V. interpreted the results and critically reviewed the manuscript: P.H.R, A.F and L.A. We exclusively used secondary and aggregated data from the public domain. Therefore, informed consent and the approval by the Research Ethics Committee are not needed, according to Resolution n. 466/2012 of the National Research Ethics Commission of the National Health Council of Brazil

## Data Availability

Data described in the manuscript, codebook, and analytic code will be made available upon request.

## Notes

### Competing Interest Statement

The authors have declared no competing interest.

### Funding Statement

This study did not receive any funding

## Reference

1. Cutland CL, Lackritz EM, Mallett-Moore T, Bardají A, Chandrasekaran R, Lahariya C, et al. Low birth weight: Case definition & guidelines for data collection, analysis, and presentation of maternal immunization safety data. Vaccine [Internet]. 2017;35(48):6492–500. Available from: https://linkinghub.elsevier.com/retrieve/pii/S0264410X17301147

2. K. C. A, Basel PL, Singh S. Low birth weight and its associated risk factors: Health facility-based case-control study. PLoS One [Internet]. 2020;15(6):e0234907. Available from: https://www.ncbi.nlm.nih.gov/pmc/articles/PMC7307746/

3. Rondó PHC, Lemos JO, Pereira JA, Oliveira JM, Innocente LR. Relationship between birthweight and arterial elasticity in childhood. Clin Sci (Lond) [Internet]. 2008 Nov [cited 2022 Nov 16];115(10):317–26. Available from: https://pubmed.ncbi.nlm.nih.gov/18393941/

4. Rondó PHC, Lemos JO, Pereira JA, Oliveira RG, Freire MBS, Sonsin PB. The relationship between birth weight and insulin resistance in childhood. Br J Nutr [Internet]. 2010 Feb [cited 2022 Nov 16];103(3):386–92. Available from: https://pubmed.ncbi.nlm.nih.gov/19772682/

5. Tela FG, Bezabih AM, Adhanu AK. Effect of pregnancy weight gain on infant birth weight among mothers attending antenatal care from private clinics in Mekelle City, Northern Ethiopia: A facility based follow-up study. PLoS One [Internet]. 2019 Mar 1 [cited 2022 Nov 15];14(3). Available from: /pmc/articles/PMC6411121/

6. K. C. A, Basel PL, Singh S. Low birth weight and its associated risk factors: Health facility-based case-control study. PLoS One [Internet]. 2020;15(6):e0234907. Available from: https://www.ncbi.nlm.nih.gov/pmc/articles/PMC7307746/

7. Pereira JA, Rondó PHC, Lemos JO, Pacheco de Souza JM, Dias RSC. The influence of birthweight on arterial blood pressure of children. Clinical Nutrition. 2010 Jun 1;29(3):337–40.

8. World Health Organization. P07 Disorders related to short gestation and low birth weight, not elsewhere classified [Internet]. nternational Classification of Diseases and Related Health Problems. 2010 [cited 2022 Nov 15]. Available from: https://scholar.google.com/scholar_lookup?title=Disorders+related+to+short+gestation+and+low+birth+weight,+not+elsewhere+classified&publication_year=2016&

9. Agbozo F, Abubakari A, Der J, Jahn A. Prevalence of low birth weight, macrosomia and stillbirth and their relationship to associated maternal risk factors in Hohoe Municipality, Ghana. Midwifery [Internet]. 2016;40:200–6. Available from: https://linkinghub.elsevier.com/retrieve/pii/S0266613816301061

10. Fan RG, Portuguez MW, Nunes ML. Cognition, behavior and social competence of preterm low birth weight children at school age. Clinics [Internet]. 2013;68(7):915–21. Available from: https://www.ncbi.nlm.nih.gov/pmc/articles/PMC3714779/

11. Mathewson KJ, Chow CHT, Dobson KG, Pope EI, Schmidt LA, van Lieshout RJ. Mental health of extremely low birth weight survivors: A systematic review and meta-analysis. Psychol Bull [Internet]. 2017 Apr 1 [cited 2022 Nov 15];143(4):347–83. Available from: https://psycnet.apa.org/journals/bul/143/4/347

12. Sicuri E, Bardají A, Sigauque B, Maixenchs M, Nhacolo A, Nhalungo D, et al. Costs Associated with Low Birth Weight in a Rural Area of Southern Mozambique. Neu J, editor. PLoS One [Internet]. 2011;6(12):e28744. Available from: https://dx.plos.org/10.1371/journal.pone.0028744

13. Barros FC, Victora CG, Matijasevich A, Santos IS, Horta BL, Silveira MF, et al. Preterm births, low birth weight, and intrauterine growth restriction in three birth cohorts in Southern Brazil: 1982, 1993 and 2004. Cad Saude Publica [Internet]. 2008 [cited 2022 Nov 15];24 Suppl 3(SUPPL.3). Available from: https://pubmed.ncbi.nlm.nih.gov/18797714/

14. de Souza Buriol VC, Hirakata V, Goldani MZ, da Silva CH. Temporal evolution of the risk factors associated with low birth weight rates in Brazilian capitals (1996-2011). Population Health Metrics 2016 14:1 [Internet]. 2016 May 3 [cited 2022 Nov 15];14(1):1–10. Available from: https://pophealthmetrics.biomedcentral.com/articles/10.1186/s12963-016-0086-0

15. Beraldo FC, Vaz IMF, Naves MMV. Nutrição, atividade física e obesidade em adultos: aspectos atuais e recomendações para prevenção e tratamento. 2022 Nov 12;14(1):57– 62. Available from: http://rmmg.org/artigo/detalhes/1521

16. Bernal JL, Cummins S, Gasparrini A. The use of controls in interrupted time series studies of public health interventions. Int J Epidemiol. 2018;47(6).

17. SINASC. Sistema de Informações sobre Nascidos Vivos (SINASC): Plataforma de Ciência de Dados aplicada à Saúde (PCDaS) | PCDaS [Internet]. Fiocruz. 2020 [cited 2022 Nov 15]. Available from: https://pcdas.icict.fiocruz.br/conjunto-de-dados/sistema-de-informacao-sobre-nascidos-vivos/

18. Lima MCB de M, Oliveira GS de, Lyra C de O, Roncalli AG, Ferreira MAF. A desigualdade espacial do Baixo Peso ao Nascer no Brasil. Cien Saude Colet [Internet]. 2013;18:2443– 52. Available from: https://www.scielosp.org/article/csc/2013.v18n8/2443-2452/

19. Jorge MHP de M, Gotlieb SLD, Laurenti R. A saúde no Brasil: análise do período 1996 a 1999. A saúde no Brasil: análise do período 1996 a 1999 [Internet]. 2001;237. Available from: https://pesquisa.bvsalud.org/portal/resource/essiqueira/biblio-935933

20. Garcia LP, Santana LR. Evolução das desigualdades socioeconômicas na mortalidade infantil no Brasil, 1993-2008. Cien Saude Colet [Internet]. 2011;16(9):3717–28. Available from: http://www.scielo.br/scielo.php?script=sci_arttext&pid=S1413-81232011001000009&lng=pt&tlng=pt

21. Pedraza DF. Sistema de informações sobre nascidos vivos: uma análise da qualidade com base na literatura. Cad Saude Colet [Internet]. 2021;29(1):143–52. Available from: http://www.scielo.br/scielo.php?script=sci_arttext&pid=S1414-462X2021000100143&tlng=pt

22. Predebon KM, Mathias TA de F, Aidar T, Rodrigues AL. Desigualdade sócio-espacial expressa por indicadores do Sistema de Informações sobre Nascidos Vivos (SINASC). Cad Saude Publica [Internet]. 2010;26(8):1583–94. Available from: http://www.scielo.br/scielo.php?script=sci_arttext&pid=S0102-311X2010000800012&lng=pt&tlng=pt

23. Silva LR da, Christoffel MM, Souza KV de. História, conquistas e perspectivas no cuidado à mulher e à criança. Texto & Contexto - Enfermagem [Internet]. 2005;14(4):585–93. Available from: http://www.scielo.br/scielo.php?script=sci_arttext&pid=S0104-07072005000400016&lng=pt&tlng=pt

24. Andrade CLT de, Szwarcwald CL, Castilho EA de. Baixo peso ao nascer no Brasil de acordo com as informações sobre nascidos vivos do Ministério da Saúde, 2005. Cad Saude Publica [Internet]. 2008;24(11):2564–72. Available from: http://www.scielo.br/scielo.php?script=sci_arttext&pid=S0102-311X2008001100011&lng=pt&tlng=pt

25. Minagawa AT, Biagoline REM, Fujimori E, de Oliveira IMV, Moreira AP de CA, Ortega LDS. Baixo peso ao nascer e condições maternas no pré-natal. Revista da Escola de Enfermagem da USP [Internet]. 2006 Dec [cited 2022 Nov 27];40(4):548–54. Available from: http://www.scielo.br/j/reeusp/a/x6cHhqW3SGFxpj94qQczG3M/?lang=pt

26. Coimbra LC, Silva AAM, Mochel EG, Alves MTSSB, Ribeiro VS, Aragão VMF, et al. Fatores associados à inadequação do uso da assistência pré-natal. Rev Saude Publica [Internet]. 2003;37:456–62. Available from: http://www.scielo.br/j/rsp/a/Jwpw8dGyCS3cGnL6JLsmYJg/

27. Gonçalves CV, Cesar JA, Mendoza-Sassi RA. Qualidade e eqüidade na assistência à gestante: um estudo de base populacional no Sul do Brasil. Cad Saude Publica [Internet]. 2009;25(11):2507–16. Available from: http://www.scielo.br/scielo.php?script=sci_arttext&pid=S0102-311X2009001100020&lng=pt&tlng=pt

28. de Albuquerque MV, Viana AL d’Ávila, de Lima LD, Ferreira MP, Fusaro ER, Iozzi FL. Regional health inequalities: changes observed in Brazil from 2000-2016. Cien Saude Colet [Internet]. 2017 Apr 1 [cited 2022 Nov 16];22(4):1055–64. Available from: http://www.scielo.br/j/csc/a/mnpHNBCXdptWTzt64rx5GSn/?lang=en

29. Igor R, Oliveira S, Aparecida M, Ferreira S. A Emenda Constitucional n° 95/2016 e as Implicações para os Recursos da Assistência Estudantil do IFRN. FINEDUCA - Revista de Financiamento da Educação [Internet]. 2022 Mar 18 [cited 2022 Nov 16];12. Available from: https://seer.ufrgs.br/index.php/fineduca/article/view/109019

30. Bahia L, Scheffer M. Financeirização na saúde. Cad Saude Publica [Internet]. 2022 Aug 26 [cited 2022 Nov 27];38(Suppl 2):e00119722. Available from: http://www.scielo.br/j/csp/a/rLvdpNdKCGsNm6bjMqcPdLj/?lang=pt

31. Brasil. Índice de Desenvolvimento Sustentável das Cidades Brasileiras [Internet]. 2022 [cited 2022 Nov 15]. Available from: https://idsc.cidadessustentaveis.org.br/map/indicators/baixo-peso-ao-nascer

