## Supplementary Material for "Social inequalities in child development: Analysis of Low-Birth-Weight trends in Brazil, 2010-2020"

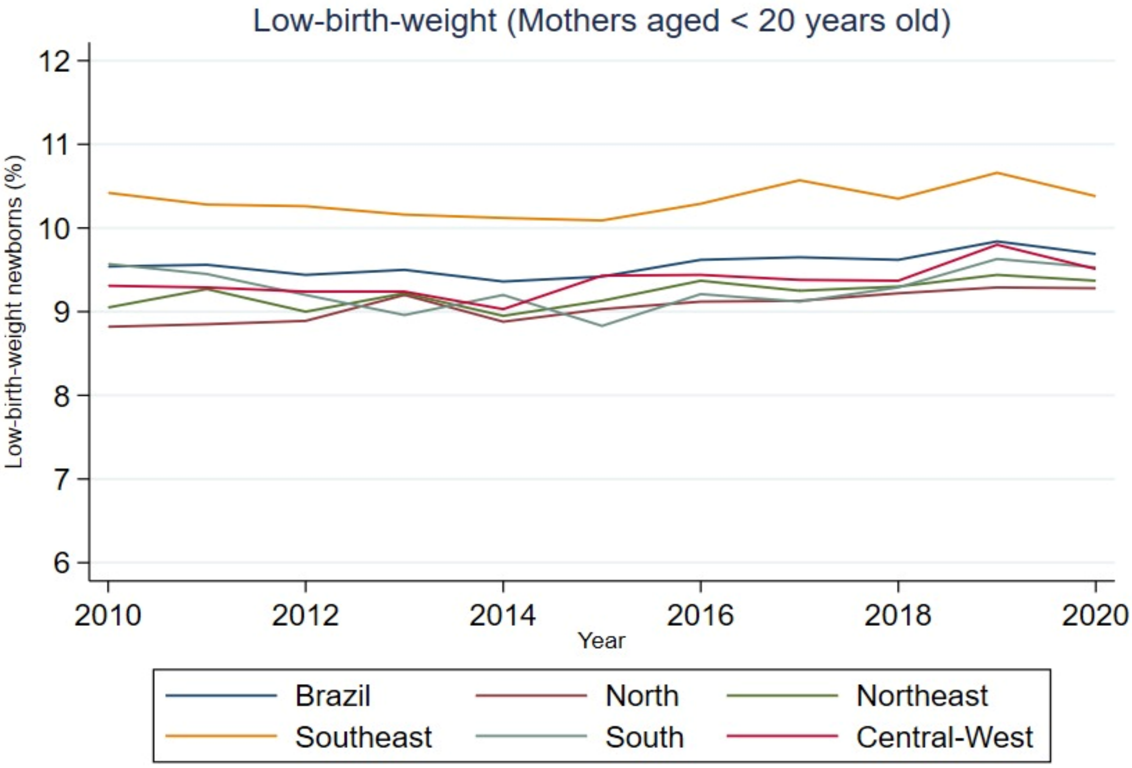


*Figure 2. Proportion of LBW (in mothers < 20 years of age) according to year of notification.*

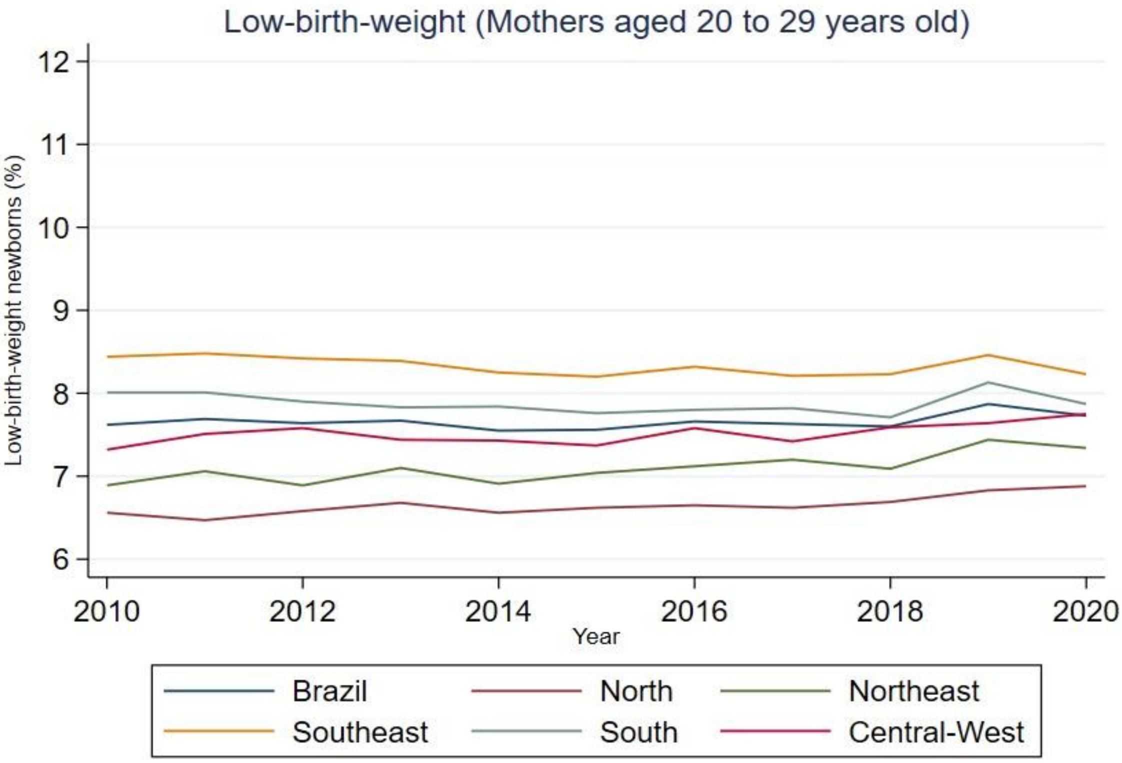


*Figure 3. Proportion of LBW (in mothers aged 20-29 years) according to year of notification.*

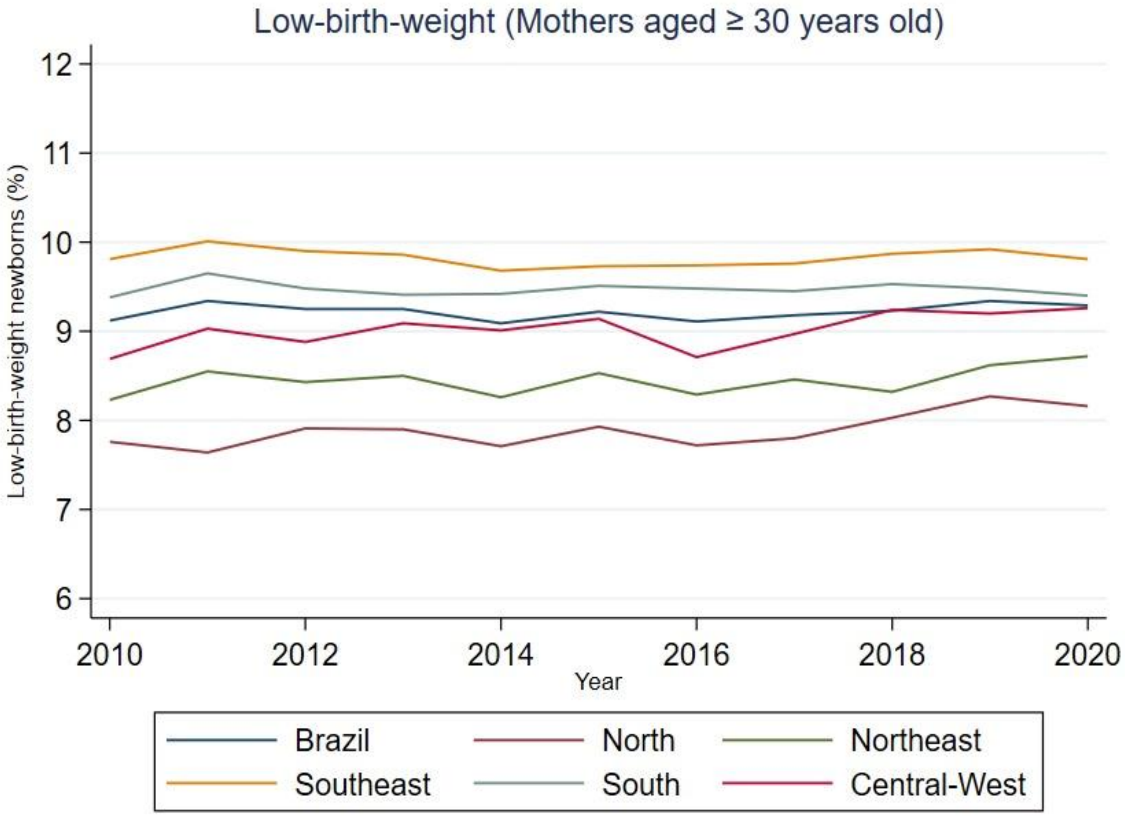


*Figure 4. proportion of LBW (in mothers ≥30 years of age) according to year of notification.*

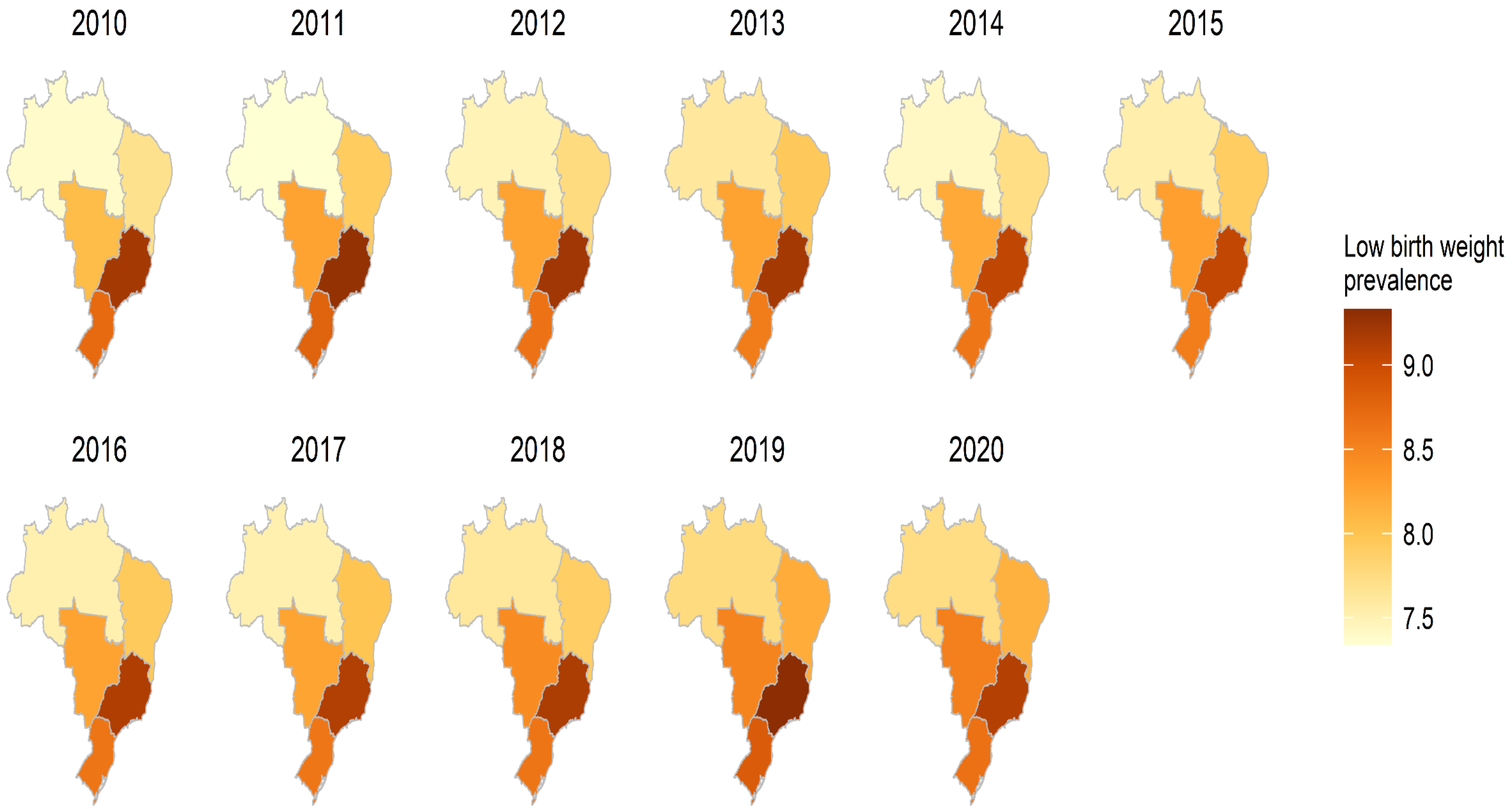

*Figure 2: Prevalence of LBW in Brazil from 2010 to 2020*
